# Sex-differential associations of BMI polygenic scores in youth with the double burden of malnutrition

**DOI:** 10.64898/2026.05.11.26352947

**Authors:** Jacus Nacis, Diana Glades Ronquillo, Michael Serafico, Reminda Bunhiyan, Mikko Glen Fernandez, Karizza Cruz, Julie Ann Jara, Josie Desnacido, Apple Joy Ducay, Eldridge Ferrer, Gerard Bryan Gonzales, Fränzel J.B. van Duijnhoven

## Abstract

**Objective:** To examine associations of BMI-related polygenic scores (PGSs) with BMI-for-age z-score (BMIz), height-for-age z-score (HAZ), and weight; assess sex-specific effects; and test PGS-by-diet interactions in youth experiencing the double burden of malnutrition.

**Methods:** In this cross-sectional study of Filipino youth aged 6-19 years, we analyzed genome-wide genotype, anthropometric, and dietary data from two 24-hour recalls. Four ancestry-standardized BMI PGSs were evaluated using linear regression adjusted for age, sex, and ancestry principal components, with platform-specific estimates combined by fixed-effects meta-analysis.

**Results:** All four PGSs were positively associated with BMIz (β range: 0.119 – 0.320). The strongest association was observed for the multi-ancestry score PGS005202 (β = 0.320; *P* = 2.39 x 10^-9^; ΔR^2^ = 4.98%). No PGS was associated with HAZ. PGS005202 and PGS005279 were associated with higher weight independent of HAZ. A significant PGS000716-by-sex interaction was observed for BMIz (q = 0.034), with an association in boys (β = 0.253; *P* = 0.002) but not in girls (β = -0.007; *P* = 0.93). No PGS-by-diet interaction remained significant after multiple-testing correction.

**Conclusions:** BMI-related PGSs were associated with adiposity-related traits, but not linear growth, in Filipino youth. Findings support sex-stratified analyses and further evaluation of ancestry-inclusive PGSs in similar pediatric settings.

## Introduction

Body mass index (BMI) is a heritable trait, with genetic factors explaining a substantial fraction of individual variation (1, 2). Genome-wide association studies (GWAS) have identified hundreds of BMI-associated loci, enabling the construction of polygenic scores (PGSs) – numerical measures that aggregate the small effects of many genetic variants to represent overall inherited susceptibility to higher body mass (2). These scores consistently predict adiposity and cardiometabolic risk from childhood through adulthood (3–5).

Yet the performance and interpretation of BMI PGSs are still uncertain outside predominantly European populations (6). This gap is particularly consequential in Southeast Asia, where populations are underrepresented in genomic research despite having distinct body composition profiles – greater adiposity at lower BMI values compared with Europeans (7). In low- and middle-income countries (LMICs) where undernutrition and obesity coexist – the double burden of malnutrition (DBM) – BMI is further complicated by linear growth deficits that inflate weight-for-height indices. Whether BMI-related PGSs in these settings reflect fat-mass variation or partly capture growth-related processes remains unresolved.

Sex may further modify the expression of genetic risk for adiposity. Differences in growth trajectories, pubertal timing, fat distribution, and hormonal regulation (8–12) may cause polygenic associations to diverge between boys and girls, yet data from pediatric LMIC populations are scarce.

Dietary intake remains a key environmental driver of adiposity and a possible modifier of genetic risk (13, 14). However, empirical evidence for gene-diet interactions in Southeast Asian children and adolescents is lacking. In populations experiencing the double burden of malnutrition, where undernutrition coexists with increasing availability of energy-dense diets, variation in dietary exposures may shape the phenotypic expression of genetic susceptibility to adiposity (7). Clarifying whether dietary exposures amplify or attenuate this susceptibility is therefore essential for interpreting genomic risk in these settings.

Here, we evaluated four established BMI polygenic scores – spanning European, East Asian, and multi-ancestry discoveries – in Filipino youth living in Metro Manila. We examined: (1) associations of PGSs with BMI-for-age z-score (BMIz), height-for-age z-score (HAZ), and weight, to distinguish adiposity-related from growth-related signals; (2) whether PGSs associations with BMIz differed by sex; and (3) whether dietary intake – total energy and macronutrients (protein, fat, and carbohydrate) – was associated with BMIz and modified PGS associations.

## Methods

### Study design and population

This study enrolled children and adolescents aged 6-19 years residing in six Metro Manila cities (Malabon, Muntinlupa, Navotas, Parañaque, Pasay, and Pasig) as participants in a genetic sub-study of the 2021 Expanded National Nutrition Survey (ENNS), a nationally representative survey conducted by the Food and Nutrition Research Institute (DOST-FNRI). Participants provided blood samples for genome-wide SNP array genotyping and were linked to ENNS anthropometric and dietary data.

The analytic sample was derived through sequential exclusions based on data completeness and predefined eligibility criteria. Participants with missing or implausible dietary intake data, those who were pregnant and lactating, and individuals without matched genotype and phenotype data were excluded. Participant selection and reasons for exclusion are summarized in Figure 1.

**Figure 1.**
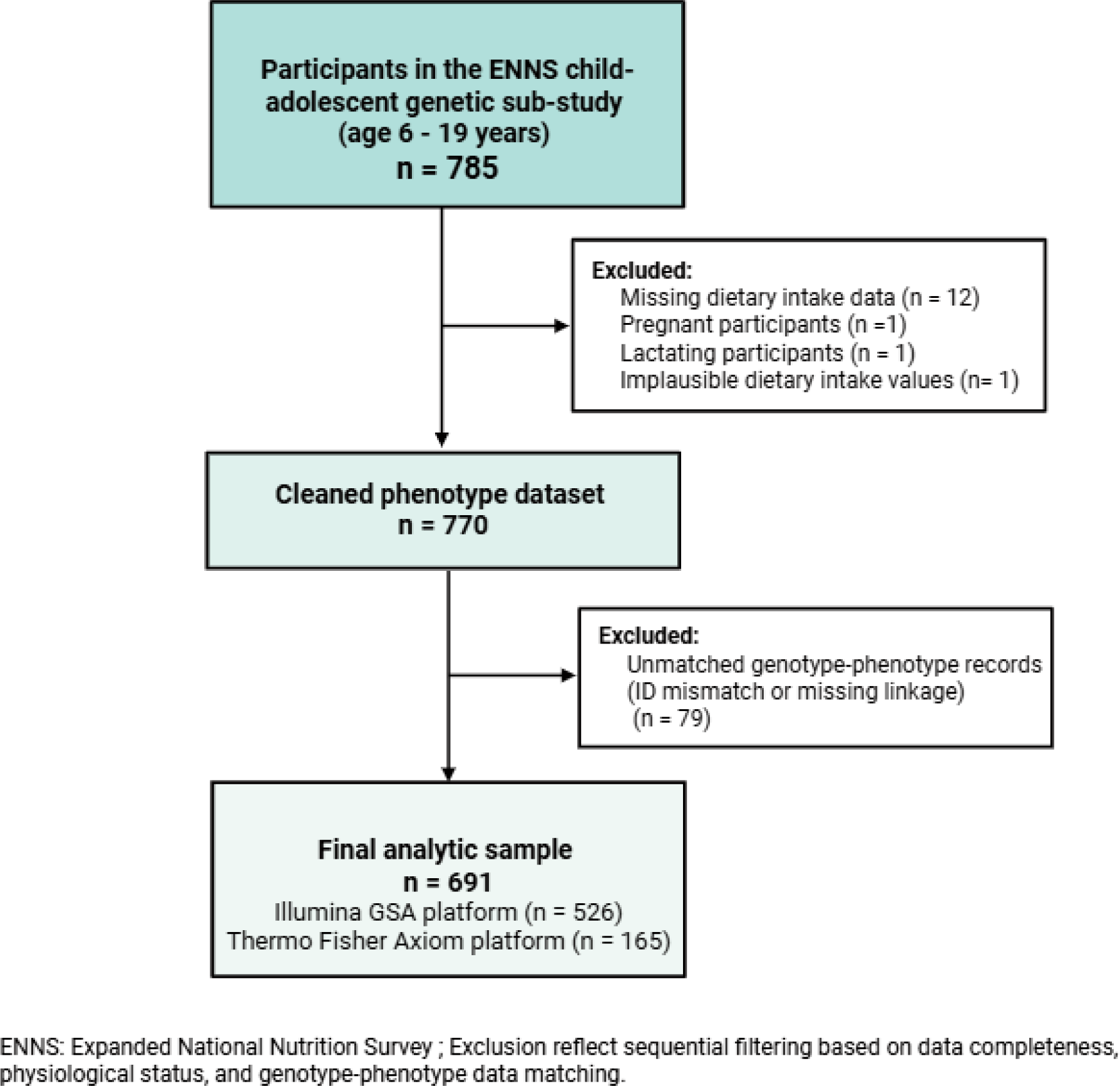
Participant selection flow diagram. Participants drawn from the ENNS child-adolescent genetic sub-study were sequentially excluded based on data completeness, physiological status, implausible dietary intake values, and genotype-phenotype mismatch, yielding a final analytic sample of 691 participants.

Written informed consent (and assent for participants aged 6-15 years) was obtained under protocols approved by the FNRI Institutional Ethics Review Committee (ENNS FIERC 2017-017; sub-study FIERC 2024-003).

### Anthropometric assessment

Weight was measured by trained field staff using a digital scale (Seca 874, Hamburg, Germany) to the nearest 0.01 kg, with duplicate readings and a third measurement obtained if the first two differed by >0.30 kg. Standing height was measured using a portable stadiometer (Seca 213, Hamburg, Germany) to the nearest 0.10 cm, with a third measurement taken if the first two differed by >0.50 cm. BMI-for-age z-score (BMIz) and height-for-age z-score (HAZ) were derived using the WHO 2007 Growth Reference for school-aged children and adolescents (5-19 years (15). WHO classification cutoffs were used for thinness, overweight/obesity, and stunting.

### Dietary assessment

Dietary intake was assessed using two-day nonconsecutive, multiple-pass 24-h food recalls administered by trained nutritionist-dietitians using standardized protocols. Household measures were standardized using measuring cups and spoons; referenced by size or number of pieces, and reported quantities were converted to gram weights using standardized conversion tables. Reported food items were matched to corresponding entries in the Philippine Food Composition Table (PhilFCT) within the Individual Dietary Evaluation System (IDES) using standardized coding procedures. Total energy intake (kcal/day) and macronutrient intakes (protein, fat, and carbohydrate in g/day) were computed as total dietary intake from all reported foods. All dietary variables were log-transformed to address skewness and approximate normality.

### Genotyping and quality control

Peripheral blood was collected onto Flinders Technology Associates (FTA) cards (Whatman, GE Healthcare, USA) and the genomic DNA was extracted using QIAamp DNA Investigator Kit (Qiagen, Germany) at the FNRI Nutritional Genomics (NuGen®) Laboratory. DNA concentration and purity were assessed spectrophotometrically, and DNA integrity was confirmed by agarose gel electrophoresis before storage at -80°C.

Genotyping was performed at accredited external laboratories using two SNP array platforms: the Illumina Infinium Global Screening Array (GSA v3.0) for the adolescent cohort and Thermo Scientific Axiom arrays for a mixed child-adolescent cohort. Standard manufacturer workflows were used, with genotype calling in GenomeStudio (Illumina) and Axiom Analysis Suite (Thermo Fisher). Platform assignment was determined solely by government procurement regulations, resulting in administrative rather than biological batch allocation.

Within each platform, genotype data underwent quality control in PLINK v1.9 (16), including harmonization of variant identifiers, integration of sex metadata, restriction to autosomal biallelic SNPs, removal of duplicate position-variants, and platform-specific variant-level filtering based on missingness and minor allele frequency (MAF < 1%). Samples with >5% missing genotypes and SNPs with >5% missingness were excluded. Eight individuals (1.2% of the cohort) exhibited discrepancies between reported and genotype-inferred sex. Given the small proportion, these individuals were retained and coded as “unknown” to avoid potential misclassification in sex-adjusted analyses. Polygenic scores were derived using autosomal SNPs only and were therefore unaffected by sex chromosome variation. After quality control filtering and linkage to phenotypic data, 526 adolescents genotyped on Illumina GSA and 165 participants (161 children, 4 adolescents) genotyped with Axiom array comprised the analytic sample. A schematic overview of the harmonized genotyping and quality control workflow across platforms is shown in Supplementary Figure S1.

### Polygenic score selection and computation

Four BMI-related polygenic scores (PGSs) were curated from the Polygenic Score Catalog (https://www.pgscatalog.org/; accessed August 2025) after screening 125 entries for phenotype relevance, discovery ancestry, variant count, and scoring file availability (17, 18). Because the study focused on pediatric outcomes, the Catalog was screened for scores derived from childhood or adolescent anthropometry; during the accession period, PGS000716 (19) was the only available score developed for early-life body size, so adult BMI scores (PGS000770, PGS005202, and PGS005279) (20–22) were retained as complementary comparators. PGS000716 (childhood body size; European ancestry), PGS000770 (adult BMI; European ancestry), PGS005202 (adult BMI; multi-ancestry), and PGS005279 (adult BMI; East Asian ancestry) were selected to represent diverse developmental contexts (childhood vs. adult) and discovery ancestries relevant to Filipino youth. Given that Filipinos occupy an intermediate position on the East-Southeast Asian genetic cline and share closer allele frequency and linkage disequilibrium patterns with East Asian than with South Asia populations (23, 24), East Asian and Asian-inclusive BMI PGSs were included. Summary characteristics of the BMI-related PGSs are shown in Supplementary Table S1.

PGSs were computed separately within each genotyping platform using the pgsc_calc pipeline (pgscatalog-utils v2.0.0) implemented in Nextflow v25.10.0 with Docker containerization (18, 25, 26). For each PGS, pgsc_calc downloaded and harmonized scoring files to the target genome build, matched variants to quality-controlled genotypes using minimum overlap thresholds (0.15 standard; 0.05 for PGS005279 on GSA due to lower SNP overlap), performed QC (variant density plots, allele matching verification), and calculated weight allele sums per participant using PLINK (16).

### PGS standardization and ancestry adjustment

Raw PGS values were first z-standardized within each platform. Kolmogorov-Smirnov tests confirmed equivalent distributions across platforms (all *P* > 0.69; Supplementary Table S2), enabling merger of platform-specific z-scores into unified predictors per PGS for pooled analyses.

Ancestry adjustment used the first 5 principal components (PCs) from LD-pruned autosomal SNPs. These PCs were projected onto the HGDP + 1000 Genomes reference panel (∼3,036 individuals) (27). Following pgsc_calc recommendations (18), a unified Python workflow aligned study genotypes to reference alleles, estimated ancestry-specific PGS means and standard deviations, and rescaled PGSs to ancestry-matched reference distributions (means = 0, SDs = 2). This transformation preserves relative ranking of individuals while removing ancestry-driven shifts in score distributions (*r* = 0.999 – 1.000 vs. raw); (Supplementary Methods 1; Supplementary Table S3).

### Statistical analysis

The primary outcome was BMI-for-age z-score (BMIz); secondary outcomes were height-for-age z-score (HAZ) and log-transformed weight. Dietary exposures included log-transformed total energy and macronutrient intakes (protein, fat, carbohydrate). All quantitative variables were analyzed as continuous measures; polygenic scores were standardized and modeled per 1-SD increment, while dietary variables and body weight were log-transformed to improve distributional assumptions. Analyses were conducted using complete-case data after exclusion of participants with missing data or implausible dietary intake. No categorical groupings of continuous variables were applied.

Linear regression models were used to evaluate the association between ancestry-standardized polygenic scores (PGS; per 1-SD increment) and BMIz, adjusted for age, sex, and ancestry principal components (PC1-PC5). Incremental variance explained (ΔR^2^) was calculated as the difference in R^2^ between models with and without the PGS, with the baseline model including age, sex, and PC1-PC5. To assess potential effect modification by sex, interaction terms (PGS x sex) were included. Sex-stratified analyses were additionally conducted to estimate associations separately in boys and girls using the same covariate set.

For secondary outcomes, the same modeling framework was applied to examine PGS in relation to HAZ and log-transformed weight. For log-transformed weight, an additional model further adjusted for HAZ to distinguish associations related to adiposity from those reflecting overall body size.

Dietary intake in relation to BMIz was evaluated using linear regression models adjusted for age, sex, and PC1-PC5. Effect modification by sex was assessed by including interaction terms between each dietary variable and sex. Meanwhile, gene-diet interactions were examined by incorporating multiplicative interaction terms (PGS x dietary variable) in BMIz models, adjusted for age, sex, and PC1-PC5. No additional energy adjustment methods (e.g., residual or density models) were applied, as total energy and macronutrient intakes were modeled directly as exposure variables.

All analyses were performed separately within each genotyping platform, and estimates were combined using fixed-effect inverse-variance meta-analysis. Between-platform heterogeneity was assessed using Cochran’s Q and I^2^ statistics.

All statistical tests were two-sided with a significance threshold of α = 0.05. *P*-values were adjusted for multiple testing using Benjamini-Hochberg false discovery rate (FDR). All analyses were conducted in SPSS version 30 (IBM Corp., Armonk, NY) and R version 2025.05.1, with meta-analyses performed using the *metafor* package.

## Results

Of the 785 eligible participants, 691 were included in the final analytic sample (Figure 1), comprising 349 girls and 342 boys. The mean age was 13.12 ± 3.77 years, mean BMIz of -0.42 ± 1.42, and HAZ of -1.02 ± 1.06. Overall, 18.7% were classified as underweight, 19.1% as stunted, 9.6% as overweight, and 7.8% as having obesity, reflecting the double burden of malnutrition in this population. Median daily energy intake was 1248 kcal (interquartile range: 927-1666 kcal), with boys reporting higher energy and macronutrient intakes than girls (Table 1).

**Table 1.**
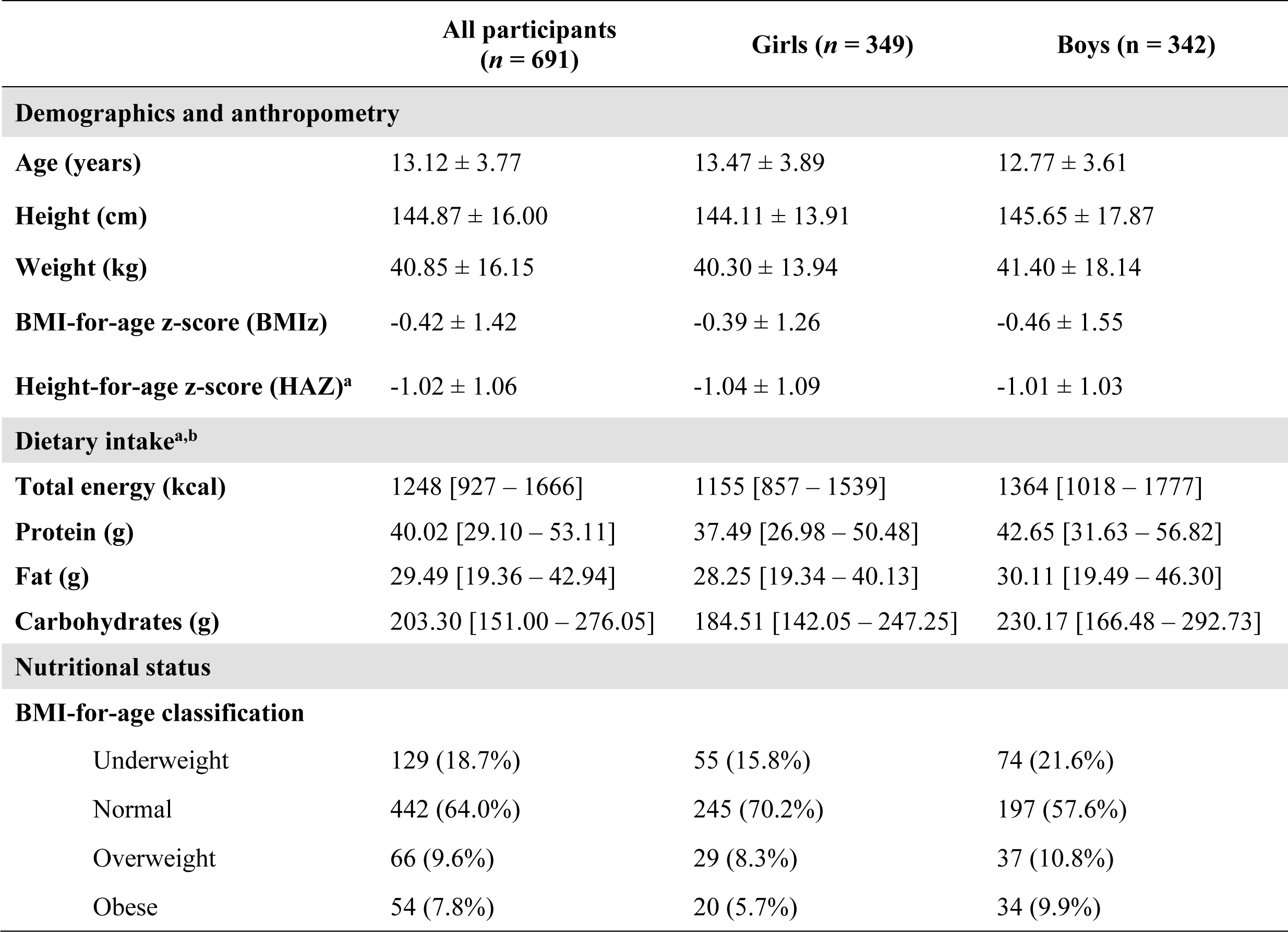
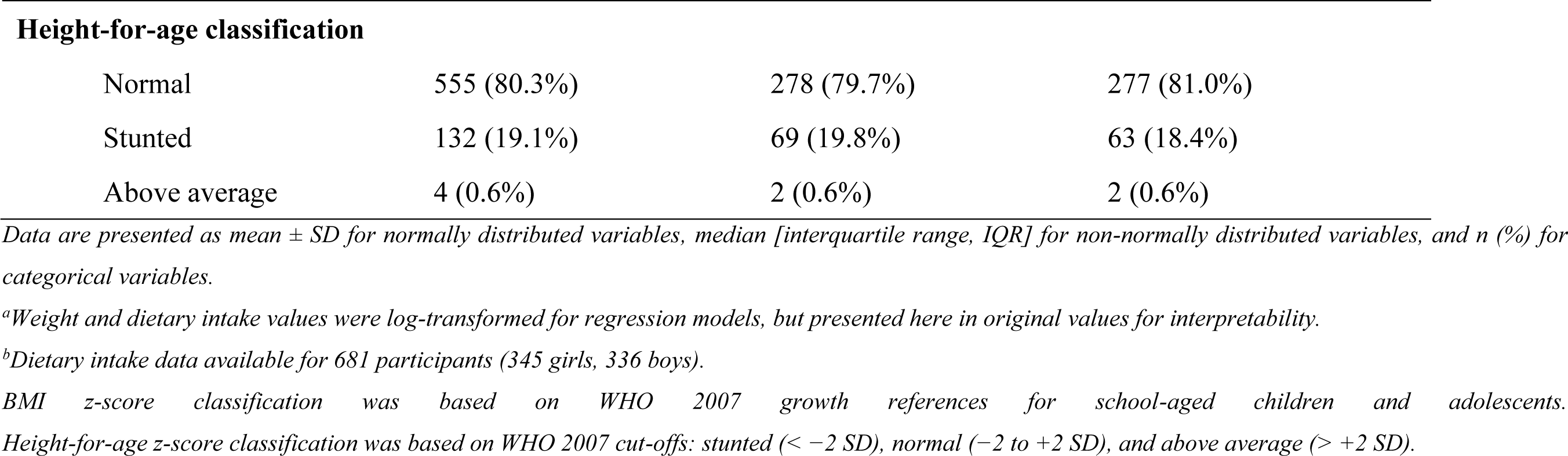
Characteristics of Filipino children and adolescents included in the analysis.

|  | **All participants**<br>**(*n* = 691)** | **Girls (*n* = 349)** | **Boys (*n* = 342)** |
| --- | --- | --- | --- |
| **Demographics and anthropometry** |  |  |  |
| **Age (years)** | 13.12 ± 3.77 | 13.47 ± 3.89 | 12.77 ± 3.61 |
| **Height (cm)** | 144.87 ± 16.00 | 144.11 ± 13.91 | 145.65 ± 17.87 |
| **Weight (kg)** | 40.85 ± 16.15 | 40.30 ± 13.94 | 41.40 ± 18.14 |
| **BMI-for-age z-score (BMIz)** | -0.42 ± 1.42 | -0.39 ± 1.26 | -0.46 ± 1.55 |
| **Height-for-age z-score (HAZ)<sup>a</sup>** | -1.02 ± 1.06 | -1.04 ± 1.09 | -1.01 ± 1.03 |
| **Dietary intake<sup>a,b</sup>** |  |  |  |
| **Total energy (kcal)** | 1248 [927 – 1666] | 1155 [857 – 1539] | 1364 [1018 – 1777] |
| **Protein (g)** | 40.02 [29.10 – 53.11] | 37.49 [26.98 – 50.48] | 42.65 [31.63 – 56.82] |
| **Fat (g)** | 29.49 [19.36 – 42.94] | 28.25 [19.34 – 40.13] | 30.11 [19.49 – 46.30] |
| **Carbohydrates (g)** | 203.30 [151.00 – 276.05] | 184.51 [142.05 – 247.25] | 230.17 [166.48 – 292.73] |
| **Nutritional status** |  |  |  |
| **BMI-for-age classification** |  |  |  |
| Underweight | 129 (18.7%) | 55 (15.8%) | 74 (21.6%) |
| Normal | 442 (64.0%) | 245 (70.2%) | 197 (57.6%) |
| Overweight | 66 (9.6%) | 29 (8.3%) | 37 (10.8%) |
| Obese | 54 (7.8%) | 20 (5.7%) | 34 (9.9%) |
| **Height-for-age classification** |  |  |  |
| Normal | 555 (80.3%) | 278 (79.7%) | 277 (81.0%) |
| Stunted | 132 (19.1%) | 69 (19.8%) | 63 (18.4%) |
| Above average | 4 (0.6%) | 2 (0.6%) | 2 (0.6%) |

*Data are presented as mean ± SD for normally distributed variables, median [interquartile range, IQR] for non-normally distributed variables, and n (%) for categorical variables.*
*<sup>a</sup>Weight and dietary intake values were log-transformed for regression models, but presented here in original values for interpretability.*
*<sup>b</sup>Dietary intake data available for 681 participants (345 girls, 336 boys).*
*BMI z-score classification was based on WHO 2007 growth references for school-aged children and adolescents.*
*Height-for-age z-score classification was based on WHO 2007 cut-offs: stunted ( $< -2$ SD), normal ( $-2$ to $+2$ SD), and above average ( $> +2$ SD).*

### Polygenic score associations with BMIz, and sex differences

All four BMI-related PGSs were significantly associated with higher BMIz (Table 2), with effect sizes ranging from β = 0.119 to 0.320. The multi-ancestry score (PGS005202) showed the strongest association (β = 0.320; P = 2.39 x 10^-9^; incremental R^2^ = 4.98%), followed by the East Asian score (PGS005279; β = 0.215; P = 6.64 x 10^-5^). All associations remained significant after false discovery rate (FDR) correction, with no between-platform heterogeneity (I^2^ = 0.0% for all BMIz models).

**Table 2.** Association of BMI-related polygenic scores with BMI-for-age z-score.

| PGS ID | Pooled $\beta$ (95% CI) | $P_{\text{FEMA}}$ | FDR q-value | $\Delta R^2$ (%) | $I^2$ (%) | Sex Int p-value | Sex Int q-value |
| --- | --- | --- | --- | --- | --- | --- | --- |
| PGS005202 | 0.320 (0.216, 0.424) | $2.39 \times 10^{-9}$ | $9.56 \times 10^{-9}$ | 4.98 | 0.0 | 0.0495* | 0.066 |
| PGS005279 | 0.215 (0.110, 0.320) | $6.64 \times 10^{-5}$ | $1.33 \times 10^{-4}$ | 2.17 | 0.0 | 0.0359* | 0.066 |
| PGS000770 | 0.150 (0.043, 0.256) | $5.97 \times 10^{-3}$ | $7.96 \times 10^{-3}$ | 0.76 | 0.0 | 0.213 | 0.213 |
| PGS000716 | 0.119 (0.010, 0.226) | $3.13 \times 10^{-2}$ | $3.13 \times 10^{-2}$ | 0.54 | 0.0 | 0.00860* | 0.034* |

*Fixed-effects meta-analyses (FEMA) results for the primary association model ( $BMI_z \sim PGS\_z + age + sex + PC1 - PC5$ ) in the pooled cohort, $n = 691$ .*
*$\Delta R^2$ (%) – incremental variance explained by adding the PGS to a baseline model including age, sex, and PC1-PC5*
*$I^2$ (%) – between platform heterogeneity; 0% indicates negligible heterogeneity between genotyping array estimates.*
*Sex Int p-value – P-value for the PGS x sex interaction term from the model $BMI_z = age + sex + PC1-PC5 + PGS + (PGS \times sex)$ .*
*Sex Int q-value –Benjamini-Hochberg FDR-adjusted P-values across the four PGS x sex interaction tests.*
*\*Indicates statistical significance after FDR correction ( $q < 0.05$ ).*

To further characterize these associations, we examined whether they differed by sex by using interaction terms (Table 2). A significant PGS x sex interaction was observed for the childhood body-size score PGS000716 (q = 0.034). Interaction terms for PGS005202 and PGS005279 were nominally significant (P < 0.05) but did not remain significant after correction for multiple testing.

Sex-stratified analyses (Table 3), showed consistently larger effect estimates in boys. For PGS000716, the association was near zero in girls (β = -0.007; *P* = 0.93) but positive in boys (β = 0.253, *P* = 0.002). Similar patterns were observed for PGS005202 (girls: β = 0.219; *P* = 0.001; boys: β = 0.452; P = 2.39 x 10^-8^) and PGS005279 (girls: β = 0.105; *P* = 0.14; boys: β = 0.352; P = 2.04 x 10^-5^).

**Table 3.**
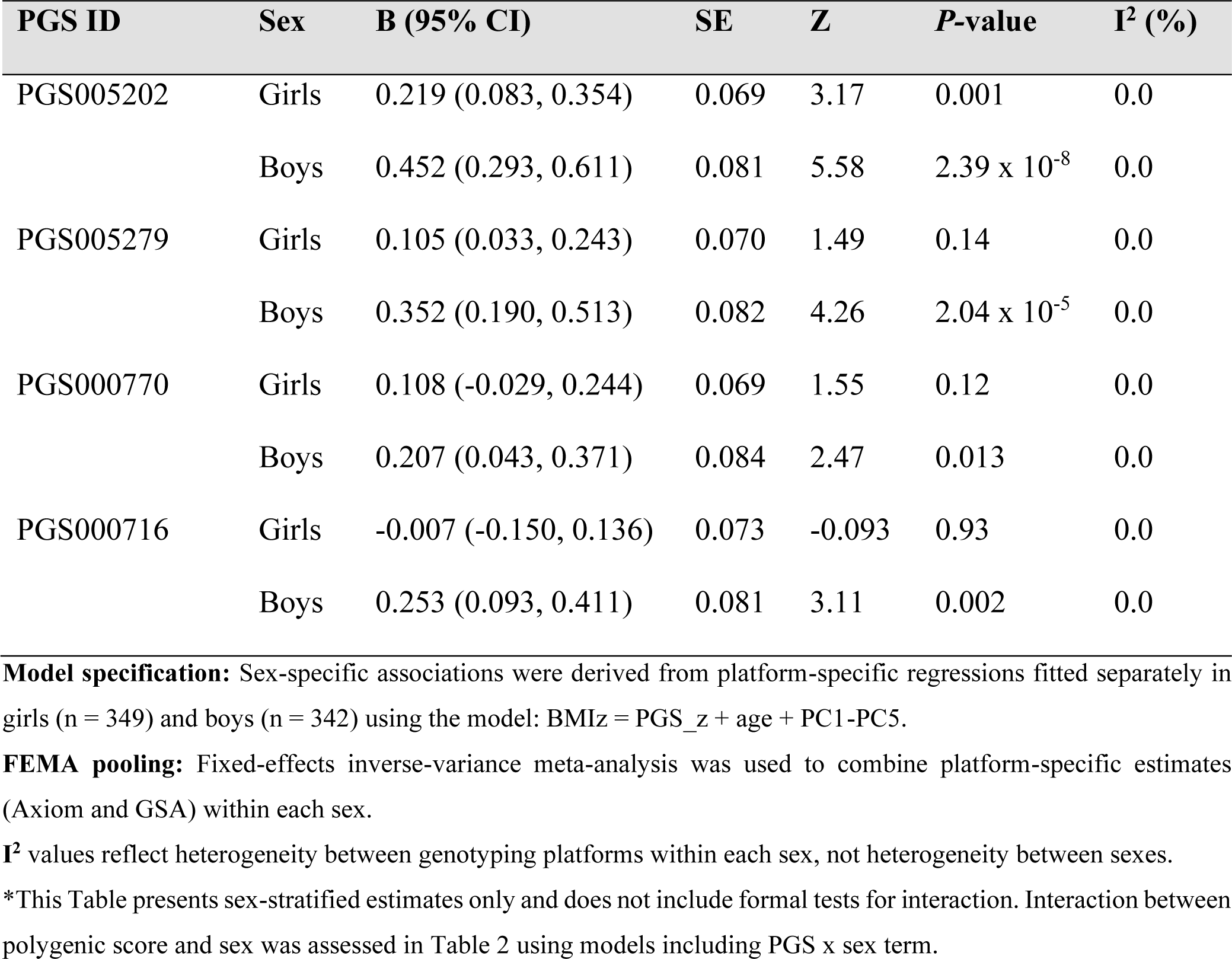
Sex-stratified associations between ancestry-normalized polygenic scores and BMI-for-age z-score (BMIz)

| PGS ID | Sex | B (95% CI) | SE | Z | P-value | I<sup>2</sup> (%) |
| --- | --- | --- | --- | --- | --- | --- |
| PGS005202 | Girls | 0.219 (0.083, 0.354) | 0.069 | 3.17 | 0.001 | 0.0 |
|  | Boys | 0.452 (0.293, 0.611) | 0.081 | 5.58 | 2.39 x 10<sup>-8</sup> | 0.0 |
| PGS005279 | Girls | 0.105 (0.033, 0.243) | 0.070 | 1.49 | 0.14 | 0.0 |
|  | Boys | 0.352 (0.190, 0.513) | 0.082 | 4.26 | 2.04 x 10<sup>-5</sup> | 0.0 |
| PGS000770 | Girls | 0.108 (-0.029, 0.244) | 0.069 | 1.55 | 0.12 | 0.0 |
|  | Boys | 0.207 (0.043, 0.371) | 0.084 | 2.47 | 0.013 | 0.0 |
| PGS000716 | Girls | -0.007 (-0.150, 0.136) | 0.073 | -0.093 | 0.93 | 0.0 |
|  | Boys | 0.253 (0.093, 0.411) | 0.081 | 3.11 | 0.002 | 0.0 |

**Model specification:** Sex-specific associations were derived from platform-specific regressions fitted separately in girls (n = 349) and boys (n = 342) using the model: BMI<sub>z</sub> = PGS<sub>z</sub> + age + PC1-PC5.
**FEMA pooling:** Fixed-effects inverse-variance meta-analysis was used to combine platform-specific estimates (Axiom and GSA) within each sex.
I<sup>2</sup> values reflect heterogeneity between genotyping platforms within each sex, not heterogeneity between sexes.
\*This Table presents sex-stratified estimates only and does not include formal tests for interaction. Interaction between polygenic score and sex was assessed in Table 2 using models including PGS x sex term.

### Polygenic score associations with weight and HAZ

Three of four PGSs (PGS005202, PGS005279, and PGS000770) were significantly associations with log-transformed weight (Table 4), with the strongest associations for PGS005202 (β = 0.048; *P* = 5.86 x 10^-3^) and PGS005279 (β = 0.036; *P* = 1.54 x 10^-4^). Further adjustments for HAZ modestly attenuated but did not materially change these estimates (PGS005202: β = 0.042; *P* = 8.07 x 10^-3^; PGS005279: β = 0.035; *P* = 1.92 x 10^-6^), indicating that weight associations were independent of linear growth. In contrast, no PGS was associated with HAZ, with all estimates near zero. Between-platform heterogeneity was negligible except for PGS000770 (I^2^ = 29.2%).

**Table 4.** Associations of BMI-related polygenic scores with height-for-age z-score (HAZ), log-transformed weight, and log-transformed weight adjusted for HAZ.

| Outcome | PGS ID | $\beta$ (95% CI) | $P_{\text{FEMA}}$ | $I^2$ (%) |
| --- | --- | --- | --- | --- |
| **Height-for-age z-score (HAZ)** | PGS005202 | -0.039 (-0.112, 0.035) | 0.042 | 0.0 |
|  | PGS005279 | -0.050 (-0.137, 0.027) | 0.214 | 0.0 |
|  | PGS000770 | 0.040 (-0.032, 0.112) | 0.278 | 0.0 |
|  | PGS000716 | 0.004 (-0.168, 0.112) | 0.917 | 0.0 |
| **Log-transformed weight** | PGS005202 | 0.048 (0.014, 0.112) | $5.86 \times 10^{-3}$ | 0.0 |
| | PGS005279 | 0.036 (0.021, 0.050) | $1.54 \times 10^{-4}$ | 0.0 |
|  | PGS000770 | 0.022 (0.003, 0.041) | 0.026 | 0.0 |
|  | PGS000716 | 0.018 (-0.002, 0.037) | 0.072 | 0.0 |
| **Log-transformed weight, adjusted for HAZ** | PGS005202 | 0.042 (0.007, 0.078) | $8.07 \times 10^{-3}$ | 0.0 |
| | PGS005279 | 0.035 (0.021, 0.050) | $1.92 \times 10^{-6}$ | 0.0 |
|  | PGS000770 | 0.020 (-0.005, 0.024) | 0.212 | 29.2 |
|  | PGS000716 | 0.014 (-0.008, 0.035) | 0.210 | 0.0 |

*Model specifications: (1) HAZ model: $HAZ = PGS\_z + age + sex + PC1-PC5$ ; (2) Log-transformed weight model: $Log(weight) = PGS\_z + age + sex + PC1-PC5$ ; (3) Log-transformed weight, adjusted for HAZ: $Log(weight) = PGS\_z + HAZ + age + sex + PC1-PC5$ .*
**$\beta$ (95% CI):** Regression coefficient per 1 SD increase in ancestry-standardized PGS.
**$P_{\text{FEMA}}$ :** Fixed-effects meta-analyses P-value.
**$I^2$ (%)** – between platform heterogeneity; 0% indicates negligible heterogeneity between genotyping array estimate

### Dietary intake associations with BMIz and modification by sex

Higher total energy, protein, and carbohydrate intakes were positively associated with BMIz, while fat intake was not (Table 5). The strongest associations were observed for carbohydrate (β = 0.472; *P* = 4.45 x 10^-4^) and total energy intake (β = 0.463; *P* = 8.85 x 10^-4^). Associations were stronger in boys than girls, with significant effect modification by sex was observed for total energy intake (*P*_interaction = 0.042; boys: β = 0.739; 95% CI: 0.306, 1.172; girls: β = 0.099; 95% CI: -0.246, 0.444). Weaker evidence of sex modification was observed for protein intake (*P*_interaction = 0.064), with no significant interactions for fat or carbohydrate intake.

**Table 5.** Associations of dietary intake with BMI-for-age z-score (BMIz), by sex.

| **Dietary variable**<br>**(log-transformed)** | **Overall**<br>**β (95% CI)** | **Girls (n = 349)**<br>**β (95% CI)** | **Boys (n = 342)**<br>**β (95% CI)** | **P_interaction** |
| --- | --- | --- | --- | --- |
| Total energy | 0.463 (0.191, 0.735) | 0.099 (-0.246, 0.444) | 0.739 (0.306, 1.172) | 0.042 |
| Total protein | 0.409 (0.174, 0.643) | 0.150 (-0.136, 0.436) | 0.684 (0.296, 1.073) | 0.064 |
| Total fat | 0.065 (-0.094, 0.223) | -0.080 (-0.279, 0.118) | 0.180 (-0.070, 0.430) | 0.115 |
| Total carbohydrates | 0.472 (0.210, 0.734) | 0.166 (-0.175, 0.507) | 0.672 (0.262, 1.083) | 0.115 |

*Main-effect estimates are derived from linear regression models for BMI-for-age z-score (BMIZ) on each dietary variable in the full sample, adjusted for age, sex, and ancestry principal components (PC1-PC5).*
*Sex-specific estimates are derived from sex-stratified linear regression models fitted separately in girls and boys, adjusted for age, sex, and ancestry principal components (PC1-PC5).*
*P\_interaction values were obtained from models including a dietary variable x sex interaction term: BMIZ ~ dietary variable + sex + (dietary variable x sex) + age + PC1-PC5.*
*β represents the change in BMIZ per 1-unit increase in log-transformed dietary intake (g/day for macronutrients: kcal/day for total energy).*
*Dietary variables were log-transformed prior to analysis.*

### Gene-diet interactions

No PGS x dietary intake interactions were statistically significant after FDR correction (Supplementary Table S4). A nominal interaction between PGS005202 and protein intake was observed (*P*_interaction = 0.042), but this did not remain significant after false discovery correction.

## Discussion

In this study of Filipino children and adolescents facing the double burden of malnutrition, BMI-related PGSs were consistently associated with BMIz across all four scores evaluated. This is consistent with prior pediatric studies showing the BMI polygenic scores predict adiposity during childhood and adolescence (3–5), and extends this evidence to a Southeast Asian population experiencing coexisting undernutrition and obesity.

Across the full cohort, BMI-related polygenic scores were associated with BMIz, with the largest effects observed for the multi-ancestry and East Asian-derived scores. Polygenic scores were also modestly associated with log-transformed weight independent of HAZ, but not with linear growth. Together, these findings suggest that BMI-related polygenic scores reflect adiposity-related variation rather than growth-related variation in this population.

Associations between BMI-related polygenic scores and BMIz were consistently stronger in boys than in girls. This pattern was most evident for the childhood body-size score PGS000716, which showed a significant PGS-by-sex interaction. Similar patterns were observed for the multi-ancestry (PGS005202) and East Asian (PGS005279) adult BMI scores, with larger effect estimates in boys in sex-stratified analyses. Previous studies have also reported sex-specific differences in the genetic contribution to adiposity across the life course, including stronger or earlier genetic effects in males and differences in contributing loci (28–31). These findings suggest that the phenotypic expression of adiposity-related genetic susceptibility may differ between boys and girls during childhood and adolescence.

The consistent direction of associations across sexes, coupled with larger effect sizes in boys, suggests that inherited susceptibility may be more strongly expressed in male youth. During childhood and adolescence, boys and girls differ in pubertal timing, hormonal regulation, lean mass accrual, and fat distribution (10, 12, 31, 32). These biological differences may influence how genetic predisposition translates into observable adiposity. However, because we lacked direct measures of pubertal stage, sex differences should be interpreted cautiously.

Dietary intake was also more strongly associated with BMIz in boys than in girls, but did not modify the relationship between polygenic scores and BMIz after correction for multiple testing. This pattern suggests that genetic susceptibility and dietary exposure contribute to adiposity through largely independent pathways. Rather than interacting, both factors appear to exert additive effects, with greater magnitude in boys. Similar observations have been reported previously, where gene-environment interactions for adiposity are often modest and inconsistent (33). In populations undergoing rapid nutrition transition, where inherited susceptibility and environmental exposures are both changing, these findings support consideration of sex as an important dimension in pediatric genetic epidemiology (34, 35), particularly in low- and middle-income settings (36).

BMI-related polygenic scores in this study primarily captured weight-related rather than linear growth variation. Although BMIz incorporates both height and weight, the persistence of associations for weight after adjustment for HAZ suggests that these scores reflect adiposity-related processes. This distinction is particularly relevant in populations with a high burden of stunting, where separating adiposity from linear growth is important. Genome-wide evidence supports this distinction, with BMI-associated loci enriched in pathways related to appetite regulation, adipogenesis, lipid storage, and energy balance (37, 38), whereas height-associated loci are linked to skeletal and endocrine developmental pathways (39). Our findings show that adiposity-related genetic signals remain detectable even in the presence of early-life growth constraints.

The stronger performance of the multi-ancestry and Asian-derived polygenic scores further underscores the importance of ancestry alignment. These scores may better capture the genetic architecture of Filipino populations, given shared allele frequencies and linkage disequilibrium with East Asian groups (23, 24). This finding reinforces the need for more diverse genomic discovery efforts, particularly in underrepresented Southeast Asian populations (40, 41).

This study has several strengths. To our knowledge, it is the first evaluation of BMI polygenic scores in children and adolescents from a setting affected by the double burden of malnutrition, demonstrating that genetic susceptibility to adiposity can be detected across nutritional extremes. The use of ancestry-standardized scores and harmonized analyses across two genotyping platforms reduces potential confounding from population structure and technical variation. Integrating genetic, anthropometric, and dietary data within a single framework also provides a broader view of adiposity during a critical developmental window.

The use of adult-derived BMI polygenic scores in a pediatric cohort is another strength of this work. Although the underlying GWAS were conducted in adults, the observed associations with BMIz suggests that the biological pathways influencing adiposity are already active during childhood and adolescence. These findings indicate that genetic influences captured by adult-derived BMI polygenic scores are detectable earlier in the life course, even if effect sizes vary across developmental stages (42, 43).

Several limitations should also be considered. Anthropometric outcomes were measured at a single time point, limiting the assessment of longitudinal trajectories of growth and adiposity. Although genetic susceptibility is present from birth, we could examine only its association with cross-sectional anthropometric indicators rather than its changing effects over time. Dietary intake was assessed using two 24-hour recalls, which may not reflect habitual intake and are subject to recall and reporting bias. Finally, given the modest sample size and possible dietary homogeneity, the study may have been underpowered to detect gene-diet interactions. The generalizability of these findings should be considered in the context of the study population. Participants were drawn from an urban Filipino cohort in a setting characterized by the double burden of malnutrition, which may limit direct extrapolation to populations with different demographic, nutritional, or epidemiological profiles. However, the consistent association between BMI-related polygenic scores and adiposity adds to growing evidence that polygenic scores are relevant in non-European populations, while also underscoring the need for further validation in diverse cohorts.

## Conclusion

In summary, BMI-related polygenic scores were associated with adiposity-related genetic predisposition among Filipino youth, with stronger expression in boys and no relationship with linear growth. Genetic susceptibility and dietary exposures appeared to contribute largely independently, although the absence of detectable gene-diet interactions should be interpreted cautiously given the complexity of dietary exposures and the limitations of short-term dietary assessment. Overall, these results suggest that genetic influences on adiposity may be more consistently detectable than gene-diet interactions in this setting. These findings support the continued evaluation of ancestry-inclusive and sex-aware genomic approaches for studying pediatric adiposity in populations experiencing the double burden of malnutrition.

## Data Availability

All data produced in the present study are available upon reasonable request to the authors.

## Author contributions

JN conceived and designed the study, oversaw its administrative and technical implementation, obtained research funding, conducted data organization, cleaning, and analyses, drafted the manuscript, and had primary responsibility for the final content; DGR and MS supervised the implementation of the study and provided operational oversight; RB, MGF, KC, and JAJ organized the datasets and biological samples and performed DNA extraction and downstream laboratory procedures; JD supervised the collection of survey data and oversaw the organization and preprocessing of dietary data; AJD and EF performed analysis and curation of survey datasets relevant to this study; GBG and FvD contributed to the conceptualization of the study, provided scientific and technical oversight, guided data analysis and interpretation, and jointly supervised the work as senior authors. All authors critically reviewed the manuscript for important intellectual content and approved the final version for submission.

## Data Availability

Data for this study are available upon request through a data transfer agreement with the Food and Nutrition Research Institute. Interested researchers may contact the corresponding author to initiate the request.

## Acknowledgements

The authors thank the Department of Science and Technology – Food and Nutrition Research Institute (DOST-FNRI) for administrative and infrastructure support for this project. The efforts of Miss Rosemarie Dumag in providing administrative supervision are greatly appreciated. We are deeply grateful to all participants of the 2021 Expanded National Nutrition Survey for their valuable time, cooperation, and willingness to contribute to scientific research. The authors also acknowledge Rachel Evangelista, Junel Trinidad, and Christian Paul Dela Torre for their assistance in sample retrieval, data collection logistics, sample extraction, and shipment. The DOST-FNRI ENNS survey team is acknowledged for their dedication in data collection. Special thanks go to Dr. Erickson Fajiculay for his assistance during the preliminary data analysis.

## References

1. Elks CE, den Hoed M, Zhao JH, Sharp SJ, Wareham NJ, Loos RJ, et al. Variability in the heritability of body mass index: a systematic review and meta-regression. Front Endocrinol (Lausanne). 2012;3:29.

2. Loos RJF, Yeo GSH. The genetics of obesity: from discovery to biology. Nat Rev Genet. 2022;23(2):120–33.

3. Jansen PR, Vos N, van Uhm J, Dekkers IA, van der Meer R, Mannens MMAM, et al. The utility of obesity polygenic risk scores from research to clinical practice: A review. Obesity Reviews. 2024;25(11):e13810.

4. Shi M, Chen W, Sun X, Bazzano LA, He J, Razavi AC, et al. Association of Genome-Wide Polygenic Risk Score for Body Mass Index With Cardiometabolic Health From Childhood Through Midlife. Circulation: Genomic and Precision Medicine. 2022;15(4):e003375.

5. Vogelezang S, Bradfield JP, Ahluwalia TS, Curtin JA, Lakka TA, Grarup N, et al. Novel loci for childhood body mass index and shared heritability with adult cardiometabolic traits. PLoS Genet. 2020;16(10):e1008718.

6. Ju D, Hui D, Hammond DA, Wonkam A, Tishkoff SA. Importance of Including Non-European Populations in Large Human Genetic Studies to Enhance Precision Medicine. Annu Rev Biomed Data Sci. 2022;5:321–39.

7. Wells JC, Sawaya AL, Wibaek R, Mwangome M, Poullas MS, Yajnik CS, et al. The double burden of malnutrition: aetiological pathways and consequences for health. Lancet. 2020;395(10217):75–88.

8. Link JC, Reue K. Genetic Basis for Sex Differences in Obesity and Lipid Metabolism. Annu Rev Nutr. 2017;37:225–45.

9. Pulit SL, Karaderi T, Lindgren CM. Sexual dimorphisms in genetic loci linked to body fat distribution. Biosci Rep. 2017;37(1).

10. Loomba-Albrecht LA, Styne DM. Effect of puberty on body composition. Curr Opin Endocrinol Diabetes Obes. 2009;16(1):10–5.

11. Li W, Liu Q, Deng X, Chen Y, Liu S, Story M. Association between Obesity and Puberty Timing: A Systematic Review and Meta-Analysis. Int J Environ Res Public Health. 2017;14(10).

12. Garnett SP, Högler W, Blades B, Baur LA, Peat J, Lee J, et al. Relation between hormones and body composition, including bone, in prepubertal children. Am J Clin Nutr. 2004;80(4):966–72.

13. Sokary S, Almaghrbi H, Bawadi H. The Interaction Between Body Mass Index Genetic Risk Score and Dietary Intake on Weight Status: A Systematic Review. Diabetes Metab Syndr Obes. 2024;17:925–41.

14. Viljakainen H, Sorlí JV, Dahlström E, Agrawal N, Portolés O, Corella D. Interaction between genetic susceptibility to obesity and food intake on BMI in Finnish school-aged children. Sci Rep. 2023;13(1):15265.

15. de Onis M, Onyango AW, Borghi E, Siyam A, Nishida C, Siekmann J. Development of a WHO growth reference for school-aged children and adolescents. Bull World Health Organ. 2007;85(9):660–7.

16. Purcell S, Neale B, Todd-Brown K, Thomas L, Ferreira MA, Bender D, et al. PLINK: a tool set for whole-genome association and population-based linkage analyses. Am J Hum Genet. 2007;81(3):559–75.

17. Lambert SA, Gil L, Jupp S, Ritchie SC, Xu Y, Buniello A, et al. The Polygenic Score Catalog as an open database for reproducibility and systematic evaluation. Nat Genet. 2021;53(4):420–5.

18. Lambert SA, Wingfield B, Gibson JT, Gil L, Ramachandran S, Yvon F, et al. Enhancing the Polygenic Score Catalog with tools for score calculation and ancestry normalization. Nat Genet. 2024;56(10):1989–94.

19. Richardson TG, Sanderson E, Elsworth B, Tilling K, Davey Smith G. Use of genetic variation to separate the effects of early and later life adiposity on disease risk: mendelian randomisation study. Bmj. 2020;369:m1203.

20. de Toro-Martín J, Guénard F, Bouchard C, Tremblay A, Pérusse L, Vohl MC. The Challenge of Stratifying Obesity: Attempts in the Quebec Family Study. Front Genet. 2019;10:994.

21. Smit RAJ, Wade KH, Hui Q, Arias JD, Yin X, Christiansen MR, et al. Polygenic prediction of body mass index and obesity through the life course and across ancestries. Nat Med. 2025;31(9):3151–68.

22. Sutoh Y, Hachiya T, Otsuka-Yamasaki Y, Komaki S, Minabe S, Ohmomo H, et al. Healthy lifestyle practice correlates with decreased obesity prevalence in individuals with high polygenic risk: TMM CommCohort study. Journal of Human Genetics. 2025;70(1):9–15.

23. Marvelle AF, Lange LA, Qin L, Wang Y, Lange EM, Adair LS, et al. Comparison of ENCODE region SNPs between Cebu Filipino and Asian HapMap samples. J Hum Genet. 2007;52(9):729–37.

24. Larena M, Sanchez-Quinto F, Sjödin P, McKenna J, Ebeo C, Reyes R, et al. Multiple migrations to the Philippines during the last 50,000 years. Proc Natl Acad Sci U S A. 2021;118(13).

25. Di Tommaso P, Chatzou M, Floden EW, Barja PP, Palumbo E, Notredame C. Nextflow enables reproducible computational workflows. Nat Biotechnol. 2017;35(4):316–9.

26. Merkel D. Docker: lightweight Linux containers for consistent development and deployment. Linux Journal. 2014;2014.

27. Auton A, Brooks LD, Durbin RM, Garrison EP, Kang HM, Korbel JO, et al. A global reference for human genetic variation. Nature. 2015;526(7571):68–74.

28. Warrington NM, Howe LD, Wu YY, Timpson NJ, Tilling K, Pennell CE, et al. Association of a body mass index genetic risk score with growth throughout childhood and adolescence. PLoS One. 2013;8(11):e79547.

29. Dubois L, Ohm Kyvik K, Girard M, Tatone-Tokuda F, Pérusse D, Hjelmborg J, et al. Genetic and environmental contributions to weight, height, and BMI from birth to 19 years of age: an international study of over 12,000 twin pairs. PLoS One. 2012;7(2):e30153.

30. Schousboe K, Willemsen G, Kyvik KO, Mortensen J, Boomsma DI, Cornes BK, et al. Sex differences in heritability of BMI: a comparative study of results from twin studies in eight countries. Twin Res. 2003;6(5):409–21.

31. Waterfield S, Richardson TG, Davey Smith G, O’Keeffe LM, Bell JA. Life course effects of genetic susceptibility to higher body size on body fat and lean mass: prospective cohort study. International Journal of Epidemiology. 2023;52(5):1377–87.

32. Silventoinen K, Jelenkovic A, Palviainen T, Dunkel L, Kaprio J. The Association Between Puberty Timing and Body Mass Index in a Longitudinal Setting: The Contribution of Genetic Factors. Behav Genet. 2022;52(3):186–94.

33. Tan PY, Moore JB, Bai L, Tang G, Gong YY. In the context of the triple burden of malnutrition: A systematic review of gene-diet interactions and nutritional status. Crit Rev Food Sci Nutr. 2024;64(11):3235–63.

34. Shah B, Tombeau Cost K, Fuller A, Birken CS, Anderson LN. Sex and gender differences in childhood obesity: contributing to the research agenda. BMJ Nutr Prev Health. 2020;3(2):387–90.

35. Ober C, Loisel DA, Gilad Y. Sex-specific genetic architecture of human disease. Nat Rev Genet. 2008;9(12):911–22.

36. Pledger SL, Ahmadizar F. Gene-environment interactions and the effect on obesity risk in low and middle-income countries: a scoping review. Frontiers in Endocrinology. 2023;Volume 14 - 2023.

37. Turcot V, Lu Y, Highland HM, Schurmann C, Justice AE, Fine RS, et al. Protein-altering variants associated with body mass index implicate pathways that control energy intake and expenditure in obesity. Nat Genet. 2018;50(1):26–41.

38. Locke AE, Kahali B, Berndt SI, Justice AE, Pers TH, Day FR, et al. Genetic studies of body mass index yield new insights for obesity biology. Nature. 2015;518(7538):197–206.

39. Robinson MR, Hemani G, Medina-Gomez C, Mezzavilla M, Esko T, Shakhbazov K, et al. Population genetic differentiation of height and body mass index across Europe. Nat Genet. 2015;47(11):1357–62.

40. Chan SH, Bylstra Y, Teo JX, Kuan JL, Bertin N, Gonzalez-Porta M, et al. Analysis of clinically relevant variants from ancestrally diverse Asian genomes. Nature Communications. 2022;13(1):6694.

41. Wong LP, Ong RT, Poh WT, Liu X, Chen P, Li R, et al. Deep whole-genome sequencing of 100 southeast Asian Malays. Am J Hum Genet. 2013;92(1):52–66.

42. Elks CE, Loos RJ, Sharp SJ, Langenberg C, Ring SM, Timpson NJ, et al. Genetic markers of adult obesity risk are associated with greater early infancy weight gain and growth. PLoS Med. 2010;7(5):e1000284.

43. Helgeland Ø, Vaudel M, Sole-Navais P, Flatley C, Juodakis J, Bacelis J, et al. Characterization of the genetic architecture of infant and early childhood body mass index. Nat Metab. 2022;4(3):344–58.

