## Supplemental Materials for "Sex-differential associations of BMI polygenic scores in youth with the double burden of malnutrition"

### List of Supplementary Materials

**Supplementary Methods:**

**Supplementary Method 1.** Principal components and ancestry adjustment ……………….. 3

**Supplementary Tables:**

**Supplementary Table S1.** Summary of BMI-related polygenic scores (PGSs) used in this study ……..………............................................................................................................ 4

**Supplementary Table S2.** Descriptive statistics and Kolmogorov-Smirnov tests for z-standardized PGSs by genotyping platforms ………………………………………………. 6

**Supplementary Table S3.** Correlations between raw, platform-standardized and ancestry-normalized polygenic scores ………………………………………………………………. 7

**Supplementary Table S4.** Gene-diet interaction effects on BMIz ………………………….................................................................................................... 8

**Supplementary Figure:**

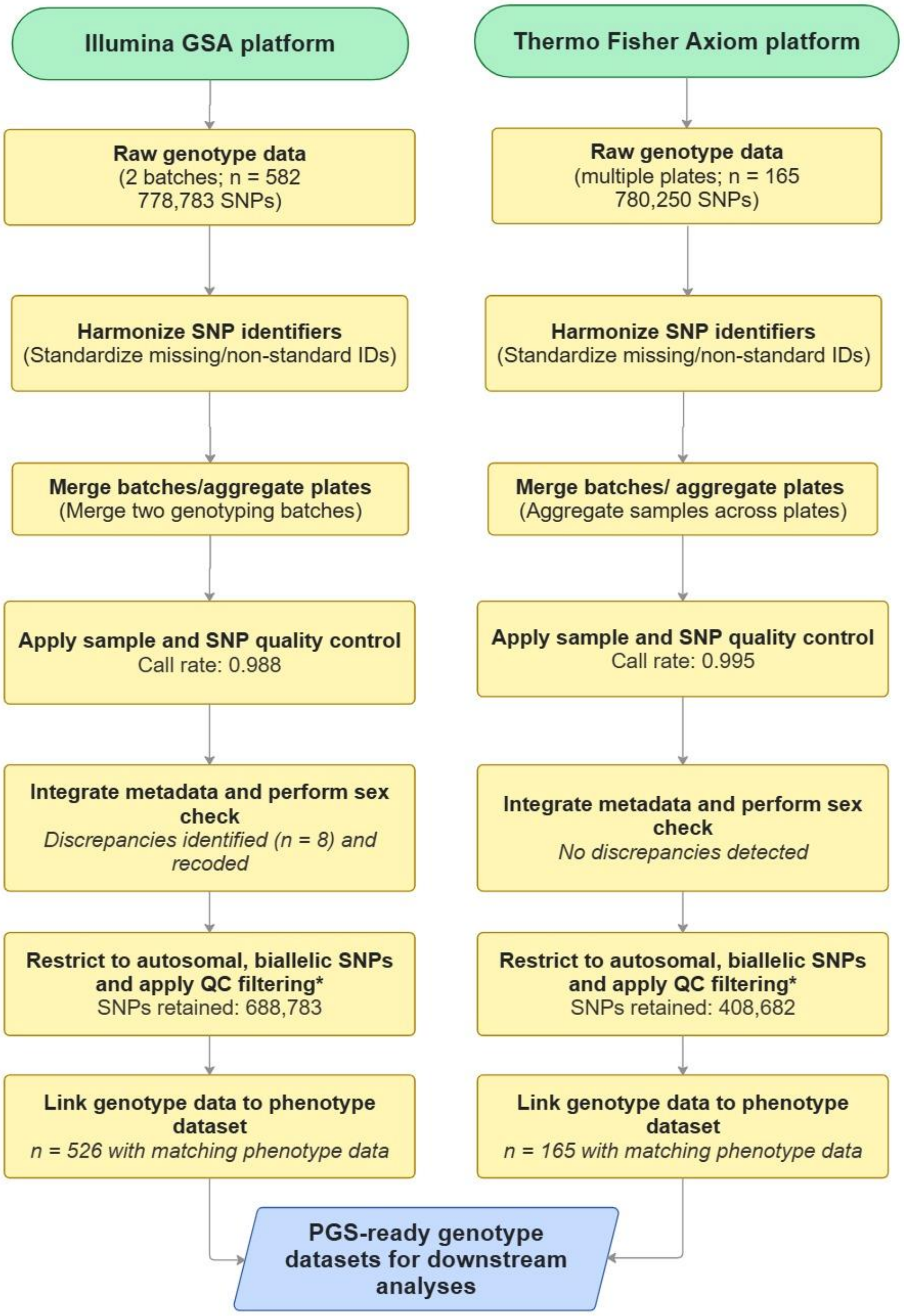


*Similar quality control steps were applied across platforms, with minor adaptations for platform-specific data structures.*

**For the Illumina GSA dataset (n =  778,783 SNPs), 36,733 SNPs were removed due to missingness, with further reductions during downstream harmonization, yielding 688,783 SNPs. For the Axiom dataset (n = 780,250 SNPs), 34,551 SNPs were removed due to missingness and 337,017 due to the minor allele frequency filtering (MAF < 1%), yielding 408,682 SNPs.*

**Supplementary Figure S1. Genotyping and quality-control workflow across platforms.** Genotype data from Illumina Global screening array and Thermo-Fisher Axiom platforms were processed using a harmonized quality control pipeline to generate polygenic score-ready datasets. All steps were conducted separately within each platform using consistent criteria, with minor adaptations to accommodate platform-specific data structures. The resulting datasets were used for downstream polygenic score analyses and cross-platform meta-analyses.

### Supplementary Method 1: Principal components and ancestry adjustment

#### *Principal component analysis of genotype data*

To characterize and adjust for population structure, principal component analysis (PCA) was performed separately within each genotype platform using LD-pruned autosomal SNPs in PLINK. For each dataset, (adolescents on GSA v3; children and some adolescents on Axiom), 20 principal components were extracted. PC1-PC2 scatterplots showed overall genetic homogeneity, with most samples forming a single dense cluster and only a few mild outliers in each cohort. Scree plots indicated that the first 5 components captured the largest share of structured variance, followed by a gradual flattening of the eigenvalue curve. Based on these patterns, the first five PCs (PC1-PC5) were retained as ancestry covariates in regression models.

#### *Ancestry normalization of polygenic scores*

To further minimize ancestry-related differences and ensure cross-platform compatibility, ancestry normalization of PGSs was performed using a Python-based unified_z workflow on a high-performance computing cluster. Study genotypes were aligned to a combined Human Genome Diversity Project (HGDP) and 1000 Genomes Project (1kGP) reference panel (~ 3,036 globally diverse individuals). For each PGS and platform, the pipeline (1) aligned study variants to the reference allele orientation, (2) estimated ancestry-weighted means and SDs of PGS values in the reference ancestry space, and (3) standardized each participant's PGS relative to these ancestry-specific parameters. This produced ancestry-normalized PGSs designed to remove ancestry-driven shifts in score distributions while preserving within-population ranking.

#### *Verification of ancestry-normalized scores*

Quality control for ancestry-normalized PGSs combined correlation analysis and visual inspection. Pearson correlations between raw platform-standardized PGSs and ancestry-normalized PGSs were close to 1.0 for all four PGS models ($r = 0.999 – 1.000$; all $p < 0.001$), indicating that the normalization preserved rank ordering while adjusting the mean and scale. These results are summarized in Supplementary Table S3.

**Supplementary Table S1.** Summary of BMI-related polygenic scores (PGSs) used in this study

| PGS ID | Reported trait/phenotype | Base GWAS/reference publication | Discovery ancestry | Variants/model type | Life-stage context | Role in this study |
|---|---|---|---|---|---|---|
| PGS005202 | BMI | Smit RAJ et al., *Nat Med* 2025 | Multi-ancestry (including East and other Asian cohorts) | ~1,022,000 variants; shrinkage-based (LDpred2-style) | Adult BMI | Asian-inclusive dense model; evaluates cross-ancestry transfer and polygenic depth in Filipino youth. |
| PGS005279 | BMI | Sutoh Y et al., *J Hum Genet* 2024 | East Asian (Tohoku TMM community cohort) | ~5,000,000 variants; LDpred-based genome-wide score | Adult BMI | Primary ancestry-matched anchor PGS; key benchmark for performance in this population. |
| PGS000770 | BMI | de Toro-Martin J et al., *Front Genet* 2020 | European | 231 genome-wide significant SNPs (GWAS-sig) | Adult BMI | European adult BMI comparator; contrasts with dense Asian- |

| | | | | | | |
|---|---|---|---|---|---|---|
| | | | | | | derived models to assess portability. |
| PGS000716 | BMI; comparative size at age 10 (self-reported) | Richardson TG, et al., *BMJ* 2020 | European (UK Biobank) | 295 genome-wide significant SNPs (GWAS-sig) | Childhood/early-life body size | Developmental comparator; tests transferability of childhood adiposity genetics. |

**Supplementary Table S2.** Descriptive statistics and Kolmogorov-Smirnov tests for z-standardized PGSs by genotyping platform

| PGS ID | Platform[a] | Mean (z) | SD | Median (z) | IQR | *P*-value[b] |
|---|---|---|---|---|---|---|
| PGS005202 | Axiom | $8.38 \times 10^{-17}$ | 1.00 | -0.117 | 1.36 | 0.726 |
| | GSA | $-3.54 \times 10^{-17}$ | 1.00 | -0.117 | 1.36 | |
| PGS005279 | Axiom | $1.98 \times 10^{-17}$ | 1.00 | -0.0827 | 1.20 | 0.836 |
| | GSA | $-4.12 \times 10^{-17}$ | 1.00 | -0.0129 | 1.26 | |
| PGS000770 | Axiom | $6.23 \times 10^{-17}$ | 1.00 | -0.117 | 1.37 | 0.986 |
| | GSA | $1.05 \times 10^{-16}$ | 1.00 | -0.117 | 1.36 | |
| PGS000716 | Axiom | $-2.57 \times 10^{-16}$ | 1.00 | -0.108 | 1.26 | 0.690 |
| | GSA | $-9.26 \times 10^{-17}$ | 1.00 | -0.102 | 1.26 | |

[a]Axiom, n = 168; GSA, n = 523.

[b]P-value from Kolmogorov-Smirnov two-sample test comparing Axiom vs GSA z-distributions for each PGS.

**Supplementary Table S3.** Correlations between raw, platform-standardized[a] and ancestry-normalized[b] polygenic scores

| PGS ID | N | Correlation (r) | *P*-value[b] |
|---|---|---|---|
| PGS005202 | 691 | 0.999 | < 0.001 |
| PGS005279 | 691 | 1.000 | < 0.001 |
| PGS000770 | 691 | 1.000 | < 0.001 |
| PGS000716 | 691 | 1.000 | < 0.001 |

[a]Raw = pre-normalized scores; [b]ancestry-adjusted = scores generated after unified z-transformation using HGDP + 1kGP ancestry reference.

**Supplementary Table S4.** Gene x dietary intake (total energy and macronutrients) interaction effects on BMIz

| PGS ID | Dietary variable (log-transformed) | *P*_interaction | *P*_FDR |
|---|---|---|---|
| PGS005202 | Total energy | 0.255 | 0.408 |
| | Total protein | 0.042 | 0.380 |
| | Total fat | 0.157 | 0.380 |
| | Total carbohydrates | 0.954 | 0.954 |
| PGS005279 | Total energy | 0.290 | 0.423 |
| | Total protein | 0.156 | 0.380 |
| | Total fat | 0.811 | 0.927 |
| | Total carbohydrates | 0.464 | 0.619 |
| PGS000770 | Total energy | 0.908 | 0.954 |
| | Total protein | 0.190 | 0.380 |
| | Total fat | 0.247 | 0.408 |
| | Total carbohydrates | 0.544 | 0.670 |
| PGS000716 | Total energy | 0.084 | 0.380 |
| | Total protein | 0.162 | 0.380 |
| | Total fat | 0.247 | 0.380 |

| | | |
|---|---|---|
| Total carbohydrates | 0.117 | 0.380 |

Models tested interaction terms between polygenic scores (PGS) and dietary variables on BMI-for-age z-score (BMIz), adjusted for age, sex, and ancestry principal components (PC1-PC5).

***P*_interaction** values were derived from Wald test of the interaction terms.

***P*_FDR:** *P*-values corrected using the Benjamini-Hochberg false discovery rate (FDR) procedure across all 16 interaction tests (4 PGS x 4 dietary exposures)
